# The influence of computer-controlled Motorbunny™ device use on sexual health and sleep behaviors

**DOI:** 10.1101/2025.04.19.25326111

**Authors:** Manus J. Donahue, Maria Garza, Ciaran Considine

## Abstract

Sleep has gained much attention as a marker of cerebral health, and it is now well-established that sleep becomes disrupted in normal aging and in the setting of neurodegeneration. However, it remains less clear how sleep behaviors can be modified by lifestyle, and specifically, whether sexual behaviors may confer sleep benefits. Evaluating these possibilities requires pilot studies that can provide necessary prerequisite data to motivate or guide larger definitive trials. Here, we utilized established Patient-Reported Outcomes Measurement Information System (PROMIS) sleep disturbance, sleep relatedness, and sexual function and satisfaction scales questionnaires of sleep behaviors and sexual satisfaction in combination with wearable actigraphy devices and performed a pilot three-way, 18-day cross-over study in adult monogamous women. Three, four-day interventions including (i) intercourse with their partner, (ii) masturbation only with a computer-controlled female vibrational device, and (iii) combined use of both the partner and the device were assessed. We tested the hypothesis that sleep efficiency and rapid-eye-movement (REM) duration were similarly increased during all forms of sexual activity compared to three-day interval washout periods of sexual abstinence. Across all participants and measures (n=4; age=34.0±10.4 years), findings were consistent with improvements in sexual satisfaction and interest in sexual activity after the trial as assessed by the PROMIS sexual satisfaction survey (t-statistic) at intake (44.6±10.5) vs. closeout (55.2±6.5). There was no significant difference in established metrics of sleep behaviors for epochs when masturbation was used vs. a partner was used; however, 75% of women exhibited evidence of an increase in sleep efficiency and REM sleep duration during the masturbation epoch compared with washout epochs with sexual abstinence, with a large effect size of 0.799. Findings are intended to provide feasibility data on the relevance of computer-controlled masturbation devices and assessing these devices with wearable technologies, and corroborate anecdotal evidence of the impact of these devices on objective markers of women’s health.

**Key points:**

- In monogamous women with no history of major neurological or psychiatric disorder, we performed a pilot (n=4) three-way, 18-day cross-over study of how self-report and actigraphy-based wellness measures depended on (i) intercourse with their partner, (ii) masturbation only with a computer-controlled female vibrational masturbation device (Motorbunny™, New Mexico, NM, USA; motorbunny.com), and (iii) combined use of both the partner and the device was applied.
- A clinical improvement, assessed by a 23.7% increase in the Patient-Reported Outcomes Measurement Information System (PROMIS) sexual function and satisfaction scales from intake to closeout was observed, consistent with improvements in self-report of sexual satisfaction over the focused sexual involvement trial.
- Sleep efficiency, defined as minutes asleep relative to minutes in bed, was most variable in washout periods of sexual abstinence, which partly stabilized for the exclusive masturbation device epoch and combined used of the masturbation device with a partner epoch. These findings are consistent with both the use of the masturbation device and combination masturbation device - partner reducing variation in sleep efficiency compared to sexual abstinence.
- The use of the masturbation device showed similar sleep efficiency and rapid eye movement (REM) sleep duration to a partner across all participants. However, in 75% of participants (i.e., 3 out of 4), REM sleep duration increased with the use of the masturbation device, with a large effect size of 0.799.

## Introduction

Sexual health is well-established as a fundamental component of population wellbeing and comprises a broad spectrum of sexual behaviors spanning reproductive care to sexual pleasure^1, 2^. Female sexual pleasure itself is a prognostic indicator of overall physical health and there are varied avenues by which such sexual pleasure can be pursued, which include sexual interactions with a partner, masturbation, or both. While not as widely evaluated in the scientific literature as other areas of sexual health, masturbation is one of the more common physical activities known to confer pleasure with ramifications extending to multiple quality of life indices. For instance, in a 2007 British national probability survey of 11,161 persons^3^, among persons aged 16 to 44 years, 95% of men and 71% of women masturbated. More importantly, these activities were noted to mitigate social pressures: 73% of men and 37% of women reported masturbating in the four weeks before a job interview and 53% of men and 18% of women reported masturbating in the previous seven days more generally^3^. These and similar large-scale population studies outline the frequency of these activities among the general population, as well as their potential relevance to social stressors.

Despite this, the health benefits of masturbation remain controversial^4-6^, and it remains unclear which benefits of inter-personal intercourse are redundant, or absent, with masturbatory activities. This issue is becoming even more fundamental as self-reported rates of masturbation have increased following the COVID-19 pandemic. Specifically, a meta-analysis of 11 studies^7^, which included a cumulative 12,350 individuals, reported a significant decrease in sexual activity in both genders (5,821 women; 3,017 men) and a significant decline in sexual function among both genders (3,974 women; 1,427 men) albeit in the presence of an increase in sexual desire and arousal. The most indicative changes in sexual behaviors during the pandemic were the increase in masturbating and usage of sex toys^7^ and it is anticipated that these activities persist to varying degrees post-pandemic. Given these statistics, it is of growing relevance to understand the impact of such activities on objective markers of health and wellbeing, although identifying such markers and quantifying these markers accurately represent ongoing challenges in the field of sexual health.

Over the past decade, sleep has gained much attention as a marker of cerebral health, partly due to the growing literature relating its relevance to neurofluid circulation and the clearance of cerebral peptides associated with aging and neurodegenerative proteinopathies, such as Alzheimer’s disease^8, 9^. There is a well-documented literature on seditious behaviors associated with extreme cases of masturbation and sleep behaviors in the setting of psychosis^6^. However, sexual behaviors, including masturbation, have also been investigated in the context of more normal sleep behaviors for more than 75 years^10-12^, with early studies showing no differences in sleep organization immediately following masturbation on polysomnographic recordings, albeit for small sample sizes and unnatural laboratory sleep monitoring^10^. Other studies have monitored sleep behaviors in natural environments, although outcomes were primarily calculated from self-report; these studies observed no gender differences in sleep outcomes following masturbation^10, 13^, with 54.1% of 284 participants in one study^13^ stating that masturbation with an orgasm improved sleep quality. In a cross-sectional study of 930 Chinese adults engaging in masturbatory activities, it was reported that sleep benefits, quantified on the self-reported Pittsburgh Sleep Quality Index (PSQI), were observed for moderate frequencies of masturbation, but reduced for extreme frequencies^14^.

While instrumental for motivating the potential impact of masturbation on sleep behaviors, the above studies suffer from the obvious caveats that overnight laboratory monitoring may not be closely related to true sleep activities, cross-sectional studies may be overly sensitive to inter-subject heterogeneity and experimental confounds, and self-report of sexual and sleep behaviors may be incompletely accurate. To address these limitations, wearable technologies, such as actigraphy devices, can be used to objectively monitor sleep behaviors in the natural environment with minimal disruption on this environment itself^15^. Additionally, the development of highly technological masturbation devices and a chronic customer base for recreation and therapeutic use^16, 17^ allows for more rigorous participant selection and experimental constrictions, thereby reducing study heterogeneity from participants with varied backgrounds, different styles of masturbation, and different levels of experience.

The overarching goal of this work was to perform a controlled pilot trial of a relatively well-characterized cohort of female participants, each with at least one year of experience with the same vibrational, computer-controlled commercially available sex device, in conjunction with objective markers of sleep behaviors from wearable actigraphy devices. We performed a three-way, 18-day cross-over study in adult women who were in a monogamous relationship for at least one year, and, three, four-day interventions including (i) intercourse with their partner, (ii) masturbation only with a computer-controlled female vibrational masturbation device, and (iii) combined use of both the partner and the device was applied. We tested the overall hypothesis that sleep behaviors are similarly increased during all forms of sexual activity compared to three-day interval washout periods of abstinence of sexual activity. The findings are intended to provide the first pilot data on how objective markers of sleep behaviors, assessed from wearable technologies, adjust during and after masturbation and should serve as a necessary prerequisite for a larger clinical trial to evaluate how different forms of sexual activity influence multiple components of sleep in a natural setting.

## Methods

### Participants and study design

Participants provided informed consent for this Institutional Review Board approved prospective study. Biological female participants working outside the sex industry with a stable monogamous partner of at least one year between the ages of 25 and 50 years were prospectively enrolled. Participants were required to be routine users of the Motorbunny (Motorbunny™, New Mexico, NM, USA; motorbunny.com) Original or Buck device for more than 12 months^18^. The Motorbunny device constituted a classic ride on top vibrator with added functionality to vibrate and rotate in a customized manner through integration with a cellular phone application. A three-way, randomized, placebo-controlled cross-over trial of Motorbunny technology was utilized. The paradigm included three separate, randomized 4-day epochs of sexual activity of (i) Motorbunny only, (ii) monogamous partner only, or (iii) combination Motorbunny and monogamous partner with a minimum of three sexual experiences per epoch (**Fig. 1**). The interval washout periods comprised three days of sexual abstinence and the cumulative participant duration was 18 days.

**Figure 1.**
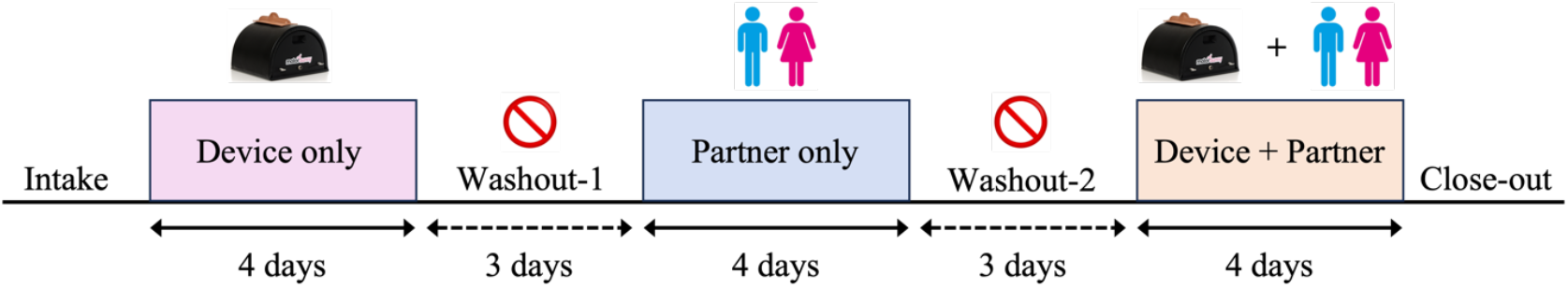
Schematic of the three-way cross-over trial design, which consisted of separate four-day epochs with use of the vibrational device only, partner only, or combined device and partner. Epochs were randomized in order across participants and included three-day interval washout periods where participants were required to abstain from all sexual activity. Sleep behaviors were monitored throughout the trial using wearable actigraphy devices, whereas established questionnaires of sleep and sexual behaviors were administered at intake and close-out.

### Surveys and questionnaires

Questionnaire sampling at transition periods of each epoch, along with contiguous actigraphy with a Fitbit Charge 6 [Google, LLC], were used to record self-reported behavioral effects and physical and sleep behavior. Questionnaires completed at the beginning and end of each intervention block were selected by a board-certified Clinical Neuropsychologist and included Hospital Anxiety and Depression Scale (HADS), PSQI, Patient-Reported Outcomes Measurement Information System (PROMIS) sleep disturbance, sleep relatedness, and sexual function and satisfaction scales, and Short-Form 36 (SF36) Social Connectedness Health. As the PROMIS sexual function and satisfaction scale queries sexual behaviors over the prior 30 days, this scale was applied at intake and at close-out to assess any changes in overall self-report of sexual behaviors before and after the structured course of sexual activity.

### Sleep behaviors

Sleep data were collected using Fitbit Charge 6 devices. Participants controlled their own Fitbit accounts via an application on their phone and authorized the collection of data without providing personally identifiable information. Monitoring began when participants enrolled and completed baseline assessments. Continuous monitoring of wear-time and battery-life was facilitated by Fitabase (San Diego, CA, USA), a comprehensive cloud-based data management platform that extracts data using the Fitbit application programming interface to generate variables. Participants on the Fitabase platform were associated with a de-identified participant identifier to ensure data privacy. The Fitbit devices provided daily summary data, including sleep duration, restless sleep time (defined by Fitbit as sleep with movement not indicating wakefulness), and sleep stages (light, deep, rapid eye movement: REM). Fitbit estimates sleep stages using a proprietary algorithm based on heart rate and movement patterns, and only estimates stages for sleep periods exceeding three hours^19^. Fitbit maps *light* sleep to N1+N2, *deep* sleep to N3, and *REM* to rapid eye movement sleep^19^. For analysis, sleep efficiency (SE) was calculated manually (minutes asleep/time in bed) to ensure data quality and prevent missing values from artificially influencing results. For sleep stage analyses, daily values were averaged across each participant’s monitoring period, using only sleep periods with available stage data. Brief sleep periods occurring during diurnal hours (i.e., 8:00 – 20:00) following cessation of the prior night’s sleep were included in that date’s total sleep calculation.

### Statistical considerations

Descriptive statistics; including mean, median, and standard deviation for continuous variables and frequency and percents for categorical variables; were calculated. As a prerequisite, a Wilcoxon rank-sum test was applied to ensure that number of sexual experiences did not differ across the three intervention epochs or between participants. To evaluate whether sexual satisfaction varied from intake to close-out (i.e., cumulative duration = 18 days), we evaluated the PROMIS sexual function and satisfaction scale. For each epoch, mean and standard deviations of measures were calculated, along with age-normed t-statistics, and effect sizes and two-sided *p*-values assessed. Here, values were interpreted in the context of population normed t-scores, with the criteria that a change in t-score of 5 or more was clinically significant, as is convention. Next, we evaluated sleep efficiency and REM sleep duration during the 4-day Motorbunny epoch relative to other epochs (e.g., partner only, Motorbunny+partner, and washout with sexual abstinence). In each case, we considered the mean as well as standard deviation of the values, with the latter providing an approximation of the variability in the sleep metric during the duration. We present the findings as both mean and standard deviation with associated p-values, percent responders (i.e., percent with an increase or decrease in the sleep metric during the epoch), and effect sizes (i.e., Cohen’s *d*). In all cases an effect size of 0.20 was interpreted as small, 0.41 as medium, 0.63 as large, and 0.87 as very large, whereas two-sided p<0.05 was considered statistically significant.

## Results

Female participants (n=4; age=34.0±10.4 years; BMI=32.2±6.6 kg/m^2^) met inclusion criteria and provided informed consent (**Table 1**). There was no significant difference (p>0.50) in the number of sexual experiences or orgasms experienced across different epochs of the cross-over trial, as was required per the trial design. Self-report assessments of sexual satisfaction; which queries degree of pleasure orgasms provided, frequency of orgasms, and interest in sexual activity over the prior 30 days; evaluated from the established categorical PROMIS assessments revealed a clinical improvement in sexual satisfaction from 44.6±10.5 (standard error = 3.1) to 55.2±6.5 (standard error = 4.1) in population normed t-score from enrollment to the conclusion of the 18-day study (**Fig. 2**; **Table 1**). These findings are consistent with improvements in sexual satisfaction and interest in sexual activity arising after focused combination use of the device and/or partner over the cumulative duration of the study.

**Table 1.** Demographic characteristics for participants, along with the intake and closeout t-scores for the PROMIS Sexual Satisfaction survey.

|  | Sex (Percent<br>Female) | Age (years) | BMI (kg/m<sup>2</sup>) | Ethnicity | Intake PROMIS Sexual<br>Satisfaction (t-score) | Closeout PROMIS Sexual<br>Satisfaction (t-score) |
| --- | --- | --- | --- | --- | --- | --- |
| *Participants* | 100 | 34.0±10.4<br>[27-49] | 32.2±6.6<br>[26.6-40.8] | 25% Hispanic<br>75% White/Caucasian | 44.6±10.5 [38.7-60.3] | 55.2±6.5 [46.7-60.3] |

**Figure 2.**
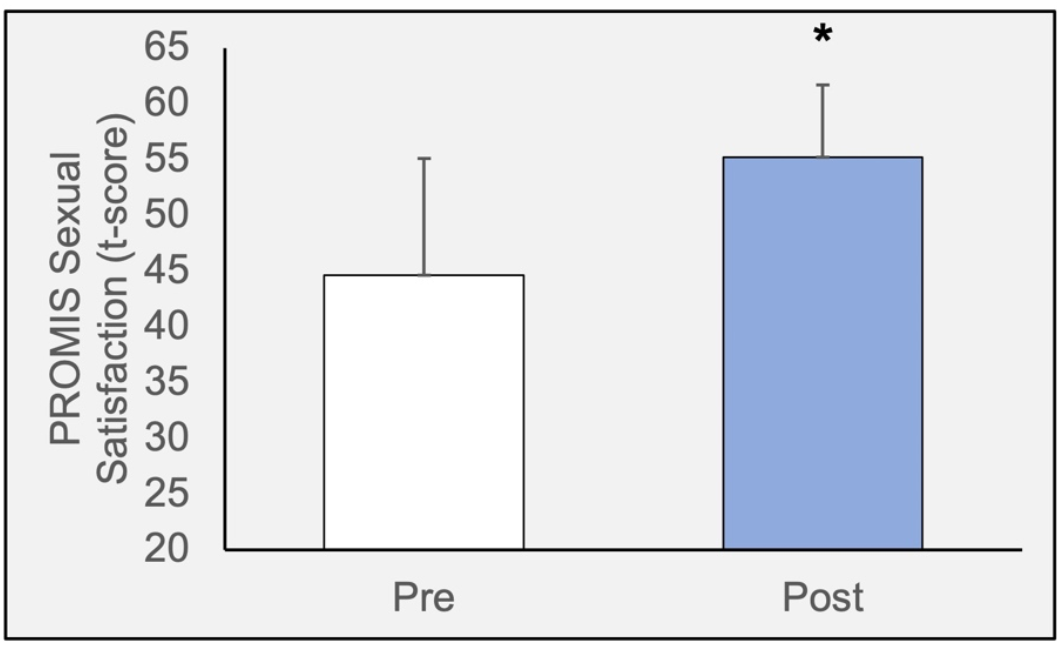
Results of the PROMIS Sexual Satisfaction survey before (pre) and after (post) the 18-day study of sexual activities. The findings reflect the cumulative effect from use of the device, partner, and device + partner. An increase in mean t-score of 10.6 was observed, consistent with a clinically significant increase in self-report of sexual satisfaction.

As self-report of prior 30-day sexual satisfaction cannot be assessed over the shorter epoch duration of four days, and as self-report is susceptible to inherent participant bias, to obtain more objective assessments of health beyond self-report, we evaluated sleep behaviors during different epochs of the cross-over trial. Missing stage data was minimal (7 nights without stage data and 13 nights with partial stage data out of 101 total nights observed across all subjects). Sleep efficiency, defined as minutes asleep relative to minutes in bed, was more variable in washout periods of sexual abstinence (standard deviation = 0.030), which partly stabilized (i.e., reduced) for the exclusive Motorbunny epoch (standard deviation = 0.017) and combined Motorbunny+partner epoch (standard deviation = 0.013) (**Table 2**). These findings are consistent with both the use of Motorbunny and Motorbunny+partner reducing variation in sleep efficiency compared to sexual abstinence.

**Table 2.** Sexual experience and sleep behavior. Summary statistics are shown for all participants (above) as well as for all participants except the participant who met criteria for Class III obesity below. Values are shown as mean ± standard deviation, with ranges for total sexual experiences in brackets.

| *All* |  |  |  |
| --- | --- | --- | --- |
|  | Sexual Experiences (count) | Sleep Efficiency (fraction) | Average REM Sleep Duration (minutes) |
| *Intake* | - | 0.912±0.022 | 101.0±37.1 |
| *Partner Only* | 2.8±1.0 [2.0-4.0] | 0.907±0.014 | 95.7±17.6 |
| *Device Only* | 3.0±0.8 [2.0-4.0] | 0.894±0.017 | 95.8±22.2 |
| *Partner+Device* | 2.8±1.0 [2.0-4.0] | 0.892±0.013 | 75.1±21.4 |
| *Washout* | 0.0±0.0 [0.0-0.0] | 0.892±0.030 | 100.1±22.7 |
| ***BMI < 40 kg/m<sup>2</sup>*** |  |  |  |
| *Intake* | - | 0.904±0.018 | 84.0±31.8 |
| *Partner Only* | 2.7±1.2 [2.0-4.0] | 0.907±0.018 | 88.7±13.0 |
| *Device Only* | 3.0±1.0 [2.0-4.0] | 0.888±0.015 | 102.7±21.2 |
| *Partner+Device* | 2.7±1.2 [2.0-4.0] | 0.898±0.009 | 67.3±17.7 |
| *Washout* | 0.0±0.0 [0.0-0.0] | 0.884±0.029 | 94.5±23.7 |

Finally, consistent with the study hypotheses, the use of Motorbunny showed similar sleep efficiency (p=0.447) and rapid eye movement (REM) sleep duration (p=0.997) to a partner across all participants. However, in 75% of participants (i.e., 3 out of 4), REM sleep duration increased with Motorbunny use with a large effect size of 0.799 (**Fig. 3**). The single participant who did not reflect this trend was noted to have Class III obesity (BMI=40.8 kg/m^2^) and the possible influence of obesity on sleep behaviors is address in the *Discussion*, and summary statistics are shown in **Table 2** with and without this participant included for completeness. It was also observed that variation in sleep measures, more so than values themselves, varied by epoch with device use generally yielding less variability in sleep measures compared to the washout periods of abstinence (**Fig. 4**). These findings are consistent with a lack of evidence to support sleep patterns improving with a partner vs. Motorbunny, and provide evidence in support of the possibility that REM sleep duration and sleep efficiency may increase following Motorbunny use, compared to sexual abstinence, as was observed in 75% of the participants.

**Figure 3.**
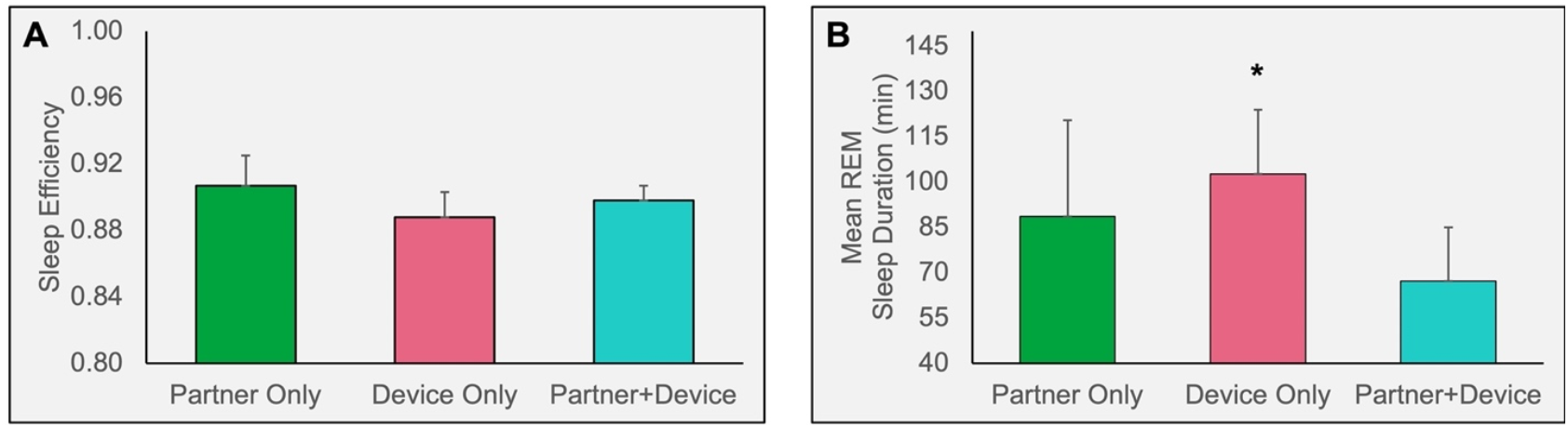
Results of the calculated assessments of (A) sleep efficiency and (B) mean rapid eye movement (REM) sleep duration for each of the interventional periods. Consistent with the primary study hypothesis, no statistically significant changes in behavior were observed for the use of the device versus the use of a partner. However, a moderate-to-large effect size (* Cohen’s d=0.799) was observed when comparing the mean REM sleep duration during the period of device use versus the partner only. Findings here exclude the participant with BMI > 40 kg/m^2^, however all quantitative values are shown in **Table 2**.

**Figure 4.**
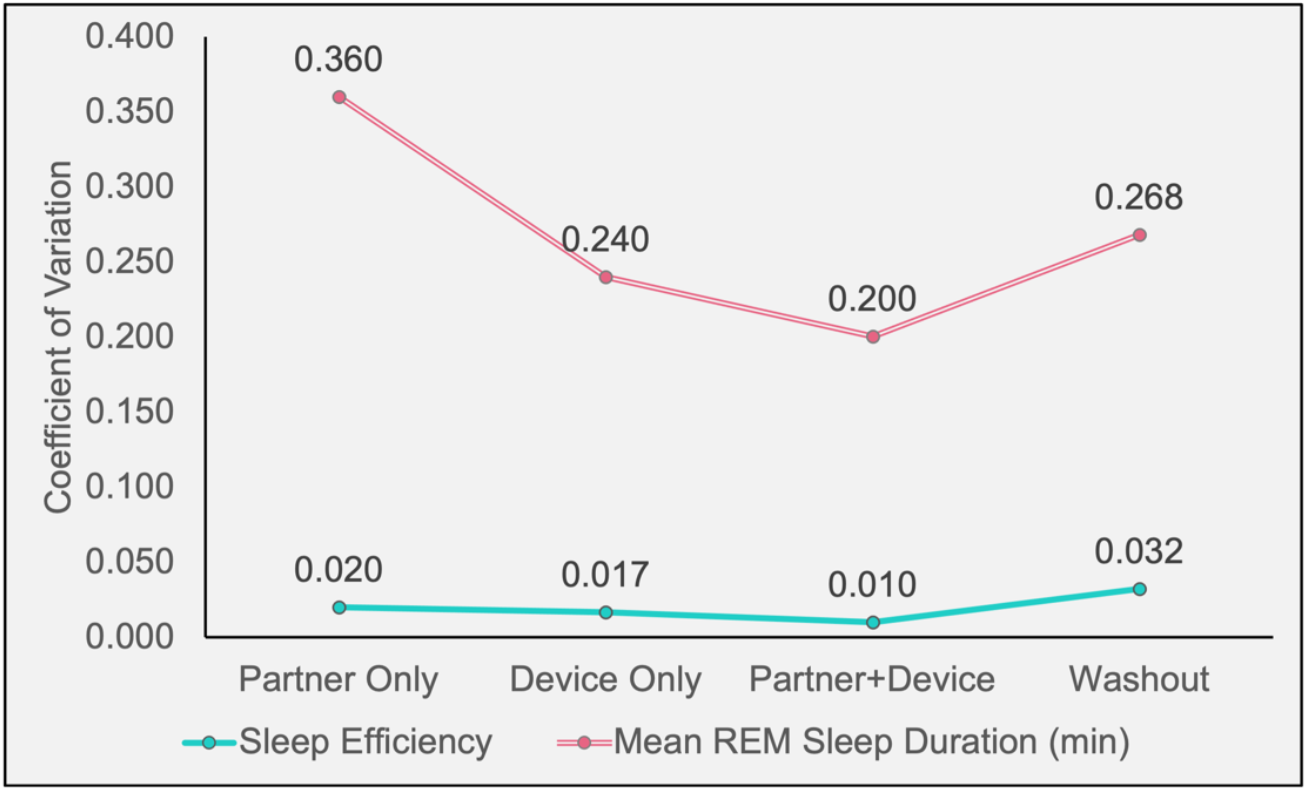
Coefficient of variation (CoV) for the different sleep measures are contrasted across different intervention epochs. Larger CoV values were observed for the mean rapid eye movement (REM) sleep duration compared to sleep efficiency. For both metrics, the CoV was larger during washout periods of sexual abstinence than for either the device use or combined partner and device use epochs. Findings are consistent with variation in sleep behaviors reducing upon device use compared to sexual abstinence.

## Discussion

We conducted a pilot, three-way, cross-over study of the possible effect of computer-controlled vibrational masturbation on female sexual satisfaction and sleep behaviors in a relatively controlled cohort of monogamous, healthy women who were chronic users of the device with no known comorbid sleep, neurological, or psychiatric conditions. The findings are consistent with improvements in sexual satisfaction and interest in sexual activity arising after focused combination use of the device and/or partner over the cumulative duration of the study. An additional finding was that there was no significant difference in established metrics of sleep behaviors for epochs when masturbation was used vs. a partner was used, despite similar sexual experiences in both cases; furthermore, 75% of women who participated exhibited evidence of an increase in sleep efficiency and REM sleep duration during the masturbation epoch compared with washout epochs with sexual abstinence. Given the early stage of wearable technologies, computer-controlled masturbation devices, and limited sample size, these findings are not definitive but provide important preliminary information that may be used to motivate larger-scale trials and corroborate anecdotal evidence of the relevance of emerging masturbation devices on women’s health.

The findings that the 18-day involvement in focused sexual activity (e.g., 12 days of sexual activity and 6 days of sexual abstinence) led to self-reported improvements in sexual wellbeing on the PROMIS assessment of sexual satisfaction is not surprising, as prior work has demonstrated improvements in sexual health, including self-reported sleep behaviors^14^, for mild masturbation frequency. The majority of studies focus on self-report, which complicates interpretation in clinical trials as (i) repeated measures administered over short periods may lead to habituation and bias, (ii) longer durations may be unnatural for many women and may introduce complications from menstration and other hormonal changes, and (iii) placebo-controlled studies are not possible in this field. As such, understanding health benefits in an objective manner will likely require pairing established sexual health surveys with objective physiological markers of health that can be monitored in a natural and comfortable setting, as attempted here.

Monitoring of sexual experiences physiologically often requires extended or overnight laboratory visits, or cumbersome and uncomfortable equipment such as EEG electrodes or invasive probes. These assessments collectively lead to metrics that may not be representative of metrics obtained in natural states. Here, we evaluated simple wearable technologies, which have been introduced in a range of other studies of health and disease, to assess sleep behaviors in a more natural setting following different sexual experiences. It was observed that inter-participant variation in sleep efficiency was 2.27-fold and 1.72-fold higher during washout sexual abstinence periods compared to combination masturbation and partner or exclusive masturbation epochs, respectively. To our knowledge, this is the first study where objective, continuous measures of activity during wake and sleep have been assessed for this technology. As sleep efficiency is well-known to deteriorate with age and is a broad measure of physical health, it is worth noting that this remained similar across different epochs and less variable compared to sexual abstinence. Similarly, REM sleep duration, which has well-documented health benefits in terms of emotional processing and memory consolidation, increased in 75% of participants. Overall, findings suggest that the computer-controlled vibrational masturbation device use provides statistically indistinguishable health benefits compared to sexual activity with a partner on the measures considered, and furthermore, exclusive use may confer health benefits relative to sexual abstinence.

Given that one participant exhibited a different trend to the group, we evaluated this participant in more detail. This participant met criteria for Class III obesity (BMI=40.8 kg/m^2^), although this was not an exclusion criterion in our original trial design. Class III obesity is associated with significant alterations in sleep-wake and circadian behaviors that can exacerbate metabolic dysfunction. The outlier’s elevated BMI may explain their unusual sleep behavior through several established mechanisms: circadian misalignment between biological rhythms and behavioral patterns^20, 21^, disrupted sleep patterns leading to increased caloric intake and reduced energy expenditure^20, 22^, impaired regulation of metabolic processes including glucose metabolism and insulin sensitivity^23-25^, dysregulation of adipokines crucial for appetite control and energy balance^26, 27^, and potentially increased sensitivity to environmental factors such as artificial light exposure that further disrupt circadian rhythms^22^. These interconnected disruptions may have affected the outlier’s sleep architecture and contributing to the observed anomalous sleep/wake behavior.

It is also worth noting that the device used in this study is a relatively complex, computer-controlled vibrational device and we selected participants who were already chronic users of this system (i.e., more than 12 months). As masturbation procedures can vary considerably across individuals, one strength of the current study design is that the same device was required in all participants. This device allows for highly customized experiences that can be controlled by a mobile phone application and participants have self-reported higher levels of sexual satisfaction from this device compared to simpler vibrational tools with more coarse settings. Future work may also benefit from understanding whether the behaviors observed here are also observed for more simple masturbation devices.

Given the above, it is worthwhile noting how these preliminary findings may be leveraged to guide future studies of sexual health. First, the study provides support that wearable technologies can be used in studies of sexual health with minimal participant discomfort, especially compared to other trial designs that require laboratory involvement or invasive monitoring. Second, inclusion criteria for such trials should be carefully designed, and it is likely that exclusion criteria should be refined to reduce confounds that may arise in this population by controlling for varied types of masturbation, hormonal changes, BMI and chronic sleep behaviors, sex industry involvement, and inter-personal relationships. We attempted to control for each of these potential confounds in this study, and as such, although the sample size of this pilot case series is relatively small, the participants are well-characterized by relationship status, device use, and overall health which reduces many confounds from larger, heterogeneous studies.

In conclusion, we conducted a pilot three-way cross-over study of the possible effect of computer-controlled vibrational masturbation on female sexual satisfaction and sleep behaviors. The findings are consistent with improvements in sexual satisfaction and interest in sexual activity arising after focused combination use of the device and/or partner over the cumulative duration of the study along with possible improvements in sleep efficiency and REM sleep duration for masturbation use compared to sexual abstinence. These findings provide important preliminary information that may be used to motivate larger-scale trials of sexual health that leverage wearable technologies.

## Data Availability

All data produced in the present study are available upon reasonable request to the authors

